# Dynamic and prognostic proteomic associations with FEV_1_ decline in chronic obstructive pulmonary disease

**DOI:** 10.1101/2024.08.07.24311507

**Authors:** Lisa Ruvuna, Kahkeshan Hijazi, Daniel E. Guzman, Claire Guo, Joseph Loureiro, Edward Khokhlovich, Melody Morris, Ma’en Obeidat, Katherine A. Pratte, Katarina M. DiLillo, Sunita Sharma, Katerina Kechris, Antonio Anzueto, Igor Barjaktarevic, Eugene R. Bleecker, Richard Casaburi, Alejandro Comellas, Christopher B. Cooper, Dawn L. DeMeo, Marilyn Foreman, Eric L. Flenaugh, MeiLan K. Han, Nicola A. Hanania, Craig P. Hersh, Jerry A. Krishnan, Wassim W. Labaki, Fernando J. Martinez, Wanda K. O’Neal, Robert Paine, Stephen P. Peters, Prescott G. Woodruff, J Michael Wells, Christine H. Wendt, Kelly B. Arnold, R. Graham Barr, Jeffrey L. Curtis, Debby Ngo, Russell P. Bowler, SPIROMICS, COPDGene and MESA-Lung Investigators

## Abstract

**Rationale:** Identification and validation of circulating biomarkers for lung function decline in COPD remains an unmet need.

**Objective:** Identify prognostic and dynamic plasma protein biomarkers of COPD progression.

**Methods:** We measured plasma proteins using SomaScan from two COPD-enriched cohorts, the Subpopulations and Intermediate Outcomes Measures in COPD Study (SPIROMICS) and Genetic Epidemiology of COPD (COPDGene), and one population-based cohort, Multi-Ethnic Study of Atherosclerosis (MESA) Lung. Using SPIROMICS as a discovery cohort, linear mixed models identified baseline proteins that predicted future change in FEV_1_ (prognostic model) and proteins whose expression changed with change in lung function (dynamic model). Findings were replicated in COPDGene and MESA-Lung. Using the COPD-enriched cohorts, Gene Set Enrichment Analysis (GSEA) identified proteins shared between COPDGene and SPIROMICS. Metascape identified significant associated pathways.

**Measurements and Main Results:** The prognostic model found 7 significant proteins in common (p < 0.05) among all 3 cohorts. After applying false discovery rate (adjusted p < 0.2), leptin remained significant in all three cohorts and growth hormone receptor remained significant in the two COPD cohorts. Elevated baseline levels of leptin and growth hormone receptor were associated with slower rate of decline in FEV_1_. Twelve proteins were nominally but not FDR significant in the dynamic model and all were distinct from the prognostic model. Metascape identified several immune related pathways unique to prognostic and dynamic proteins.

**Conclusion:** We identified leptin as the most reproducible COPD progression biomarker. The difference between prognostic and dynamic proteins suggests disease activity signatures may be different from prognosis signatures.

## INTRODUCTION

Chronic obstructive pulmonary disease (COPD) has higher mortality in individuals with rapidly progressive disease (1, 2). The gold standard for COPD diagnosis is post bronchodilator (BD) spirometric forced expiratory volume at one second (FEV_1_)/forced vital capacity (FVC) < 0.7; FEV_1_ decline is the best studied parameter to assess disease progression. Beyond continued smoking and repeated exacerbations, the factors leading to COPD progression are poorly understood (3, 4). Thus, identification of biomarkers that can be used for COPD prognosis or as pharmaceutical targets is a major unmet research need.

Although COPD affects the lungs, it has significant systemic extra-pulmonary manifestations such as muscle loss, cardiovascular disease, and osteoporosis (5, 6). Circulating (blood) biomarkers, which are less invasive to obtain than lung biomarkers, may therefore act as surrogate markers for assessing COPD risk and disease activity. While there have been multiple studies investigating cross sectional blood biomarkers for COPD (diagnostic biomarkers) in large, well-phenotyped cohorts such as ECLIPSE, SPIROMICS, and COPDGene, there are fewer studies that have investigated biomarkers of COPD progression (prognostic biomarkers) and none that have comprehensively evaluated disease activity by correlating change in biomarker concentration to disease progression (dynamic biomarkers) (7). A major limitation to many biomarker publications is that they lack external validation or do not replicate across different publications, possibly due to cohort heterogeneity, e.g., population-based versus enriched for COPD (8).

There are several reasons why a large-scale, longitudinal biomarker study with validation in independent diverse cohorts is the best approach to identify COPD progression biomarkers. First, COPD is a heterogenous condition and there are likely a plethora of multifactorial pathologic changes that lead to airflow obstruction over time. Second, smaller studies have shown that effect sizes of individual protein biomarker of COPD are small, so a comprehensive approach including combinations of biomarkers is most likely to improve modeling of COPD progression (9). Third, most biomarker studies include predominantly European ancestry subjects, and it is not clear whether findings can be replicated in non-European ancestry subjects, limiting the generalizability of the findings. Fourth, COPD is a dynamic disease with intermittent progression. Thus, disease activity biomarkers are best assessed as biomarkers that change as the disease progresses. Identifying prognostic and dynamic biomarkers of FEV_1_ decline may provide better insight into COPD progression.

This study utilizes an innovative proteomic platform (SomaScan) to identify prognostic and dynamic proteins associated with decline in FEV_1_ across three independent cohorts, with the Subpopulations and Intermediate Outcomes Measures in COPD Study (SPIROMICS) as the discovery cohort, and the Genetic Epidemiology of COPD (COPDGene) and Multi-Ethnic Study of Atherosclerosis (MESA) Lung Study as the validation cohorts.

## METHODS

### Cohort Descriptions

We analyzed three NIH-funded, multi-center observational cohort studies **(Figure 1)** all of which were approved by local Institutional Review Boards. All participants provided informed written consent. The full protocols for participant enrollment for each cohort were previously described elsewhere (10,12–14).

**Figure 1:**
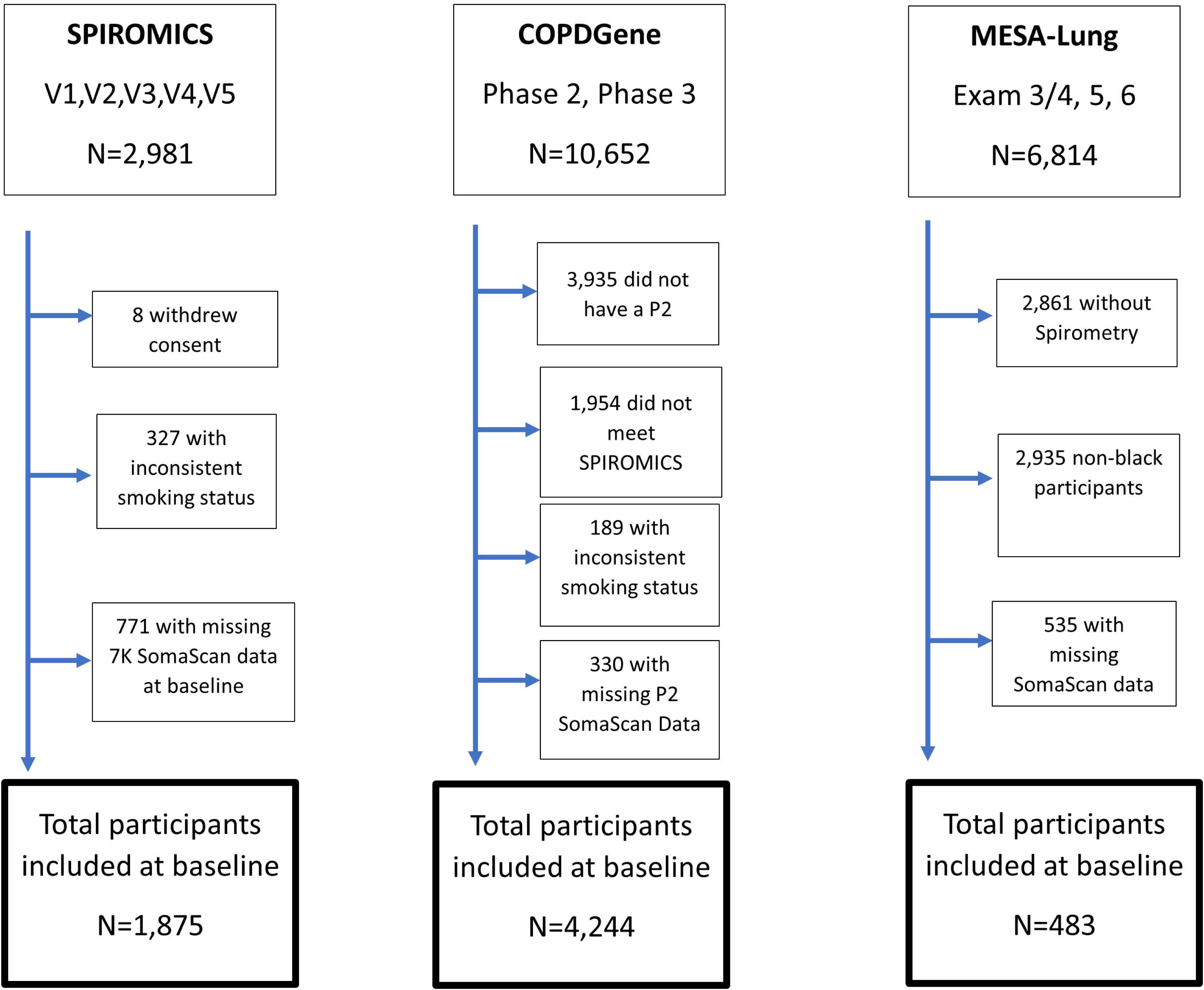
Consort Diagram. Flow diagram depicting the number of participants included after application of our study specific exclusion criteria. The SPIROMICS enrollment criteria was applied to the COPDGene cohort to reduce the variability between participants in each COPD-enriched cohort.

From 2010 to 2015 SPIROMICS <u>(ClinialTrials.gov Identifier: NCT01969344</u>) enrolled 2,981 participants ages 40-80 years old into 4 strata: never smokers (< 1 pack year), smokers with normal spirometry, mild/moderate COPD, and severe COPD. Blood specimens and spirometry were collected at baseline, year 1, year 3 and a final visit that occurred approximately 5-7 years after the baseline visit.

From 2013 to 2017, COPDGene (ClinialTrials.gov Identifier: NCT00608764), enrolled 10,198 non-Hispanic white and non-Hispanic Black never smokers, former smokers, and current smokers ages 45-80 with and without COPD with follow up visits occurring 5 years and 10 years from their initial visit.

From 2002 to 2004, MESA <u>(ClinialTrials.gov Identifier: NCT00005487)</u> recruited a multi-ethnic cohort of 6,814 participants from the community who self-identified as either White, Black, Hispanic, or Asian race/ethnicity, were ages 45-84 years old and free of clinical cardiovascular disease to investigate the prevalence, correlates, and progression of subclinical cardiovascular disease (15). From 2004 to 2006, the MESA-Lung Study enrolled 3,965 MESA participants who were sampled randomly among those who consented to genetic analysis, underwent baseline measures of endothelial function, and attended an examination during the recruitment period (16). The current replication sample was limited to 483 Black individuals who had at least 1 spirometry measurement over up to 14 years of follow-up.

### Biomarker Panel

Plasma samples were assayed on aptamer-based SomaScan platforms Version 4.1 (7,288 human SOMAmer that map to 6,467 unique proteins) in SPIROMICS and MESA-Lung and version 4.0 (4,979 human SOMAmers that map to 4,860 unique proteins) in COPDGene (10). Additional information can be found in the **<u>supplemental text</u>.**

### Clinical Definitions and Study Population

COPD was defined by post-bronchodilator FEV_1_ to FVC ratio < 0.70; in MESA-Lung, only pre-bronchodilator spirometry measurements were available in Exam 3 and thus used for Exams 3 through 6 to define COPD. All three cohorts used self-defined race, chosen from a limited number of categories. We refer to the selected choices of “Black” and/or “African American” as “Black” and where participants selected “White” and/or the historically used category “Caucasian” we refer to their race/ancestry as “White.”

All participants who consented to genetic analysis, had available proteomics results, and spirometry for at least 1 visit were included **(Figure 1).** To better compare between COPDGene and SPIROMICS, COPDGene subjects were categorized into the 4 SPIROMICS enrollment strata and excluded if they did not meet strata criteria. We minimized variability caused by smoking in SPIROMICS and COPDGene by analyzing subjects who maintained consistent smoking status throughout the study. Since MESA-Lung is a population cohort and had a smaller sample size, we did not limit to those who maintained consistent smoking status and we did not apply the SPIROMICS enrollment criteria. SomaScan v4.0 results were only run at the 5 and 10 year follow up for COPDGene. Thus, we included only participants who had at least 1 visit at the 5 or 10 years follow up with spirometry and proteomic results.

The final datasets consisted of N=1,875 subjects for SPIROMICS, N=4,244 for COPDGene and N=483 for MESA-Lung **(Figure 1).**

### Statistical Analysis

To identify baseline prognostic biomarkers of disease progression, we used a linear mixed effects model to identify proteins at baseline associated with change in FEV_1_ over the follow-up period (fixed effect of protein*time interaction). Clinical covariates modeled as fixed effects included age, age^2^, race, sex, smoking status, time-varying pack years, exacerbation history prior to baseline, baseline FVC, and time (in years) since baseline (first visit with protein measurement) in all 3 cohorts. A Benjamini-Hochberg adjusted p-value < 0.20 denoted false discovery rate (FDR) significant results and a p-value < 0.05 was used for nominal significance.

To assess multi-protein prognostic model performance, we examined the Akaike information criteria (AIC), Bayesian information criteria (BIC), and marginal R2 when each identified protein was added to the base clinical model individually for the COPD-enriched cohorts only. A backward selection approach was then applied to identify a combination of proteins that best improved the clinical model for with SPIROMICS as discovery and COPDGene as the validation cohort.

To characterize the dynamic protein level changes associated with lung function change over time, we used a linear mixed effect model to examine FEV_1_ versus the interaction of longitudinal protein levels and time in years (protein*years interaction) for SPIROMICS and COPDGene. This model was not assessed in MESA-Lung as only baseline SomaScan samples were available. Covariates included age, body mass index (BMI), smoking status, smoking pack-years, sex, and race. FDR and nominal significance were defined as they were for the prognostic model.

### Common Enrichment Pathways for FEV_1_ Decline

We used Gene Set Enrichment Analysis (GSEA) (v4.2.3) to identify the top protein relationships across SPIROMICS and COPDGene from the prognostic and dynamic change models, respectively (17, 18). GSEA analyzed the top 200 proteins associated with change in FEV_1_ in COPDGene and all 7k proteins ranked by t-statistics for FEV_1_ decline from both prognostic and dynamic models. Significant enrichment was defined by an FDR corrected p-value < 0.05. Most strongly associated leading edge proteins across the two cohorts in both models were used in Metascape functional enrichment analysis (19). A minimum enrichment of 1.5 was required to be included and p-value < 0.05 was used to identify statistically significant pathways.

Additional information on methods are provided under the supplemental text.

## RESULTS

### Characteristics of study population

The baseline characteristics of cohort participants are described in **Table 1**. As a population-based cohort, MESA-Lung participants had fewer cumulative smoking pack years, less prevalent COPD, and better lung function. The total analyzed follow-up periods of the three cohorts were similar (∼5 years).

**Table 1:** Cohort Characteristics.

|  | **SPIROMICS**<br>**(N=1,875)** | **COPDGene**<br>**(N=4,244)** | **MESA-Lung**<br>**(N = 483)** |
| --- | --- | --- | --- |
| **Age (years) Mean (SD)** | 63.6 (9.09) | 65.6 (8.7) | 68.5 (9.3) |
| **Race** |  |  |  |
| White | 1,462 (78.0%) | 3,092 (72.9%) | 0 (0%) |
| Black | 329 (17.5%) | 1,152 (27.1%) | 483 (100%) |
| Other | 84 (4.5%) | 0 (0%) | 0 (0%) |
| **Gender or Sex** |  |  |  |
| Male | 995 (53.1%) | 2,184 (51.5%) | 225 (46.6%) |
| Female | 880 (46.9%) | 2,060 (48.5%) | 226 (46.8%) |
| **Smoking Status** |  |  |  |
| Consistent former smoker | 1,118 (59.6%) | 2,380 (56.1%) | 215 (44.5%) |
| Consistent never smoker | 144 (7.7%) | 350 (8.2%) | 218 (45.1%) |
| Consistent smoker | 611 (32.6%) | 1,514 (35.7%) | 50 (10.4%) |
| **Smoking pack-years (years) Median (5<sup>th</sup> and 95<sup>th</sup> percentile)** | 42.0 (0, 95.0) | 41.4 (0, 90.0) | 14.5 (0.75, 57.0) |
| **Number of exacerbations treated with antibiotics or steroids in 12 months before baseline visit Mean (SD)** | 0.3 (0.854) | 0.3 (0.811) | NA** |
| **Post bronchodilator FEV<sub>1</sub> (liters) Mean (SD)** | 2.6 (0.932) | 2.2 (0.897) | 2.2 (0.6)* |
| **% predicted post bronchodilator FEV<sub>1</sub> Mean (SD)** | 75.2 (27.1) | 79.0 (26.2) | 95.5 (20.3)* |
| **Observed post bronchodilator FVC (liters) Mean (SD)** | 3.5 (1.03) | 3.2 (0.988) | 2.9 (0.8)* |
| **% predicted post bronchodilator FVC Mean (SD)** | 92.0 (18.2) | 89.1 (17.8) | 98.0 (20.2) |

Definition of abbreviations: SPIROMICS = Subpopulations and Intermediate Outcomes Measures in COPD Study; COPDGene = Genetic Epidemiology of COPD; MESA-Lung = Multi-Ethnic Study of Atherosclerosis-Lung
\*pre-bronchodilator spirometry value
\*\*Exacerbation history not obtained

### Prognostic Protein Biomarkers for changes in FEV_1_

The prognostic model focused on identifying baseline proteins that can predict future changes in FEV_1_. In the SPIROMICS (discovery) cohort, we identified 365 SOMAmers representing 347 proteins associated with FEV_1_ decline at nominal statistical significance (p-value < 0.05); 31 and 25 respectively remained statistically significant after FDR adjustments (adjusted p-value < 0.2) (Table E1). Next, we sought to see which significant proteins from the SPIROMICS analysis were also significant in a similar COPD-enriched cohort (COPDGene) and a population cohort of Black Americans (MESA-Lung) **(Figure 2A)**. In COPDGene, there were 406 SOMAmers representing 401 proteins that were also significantly associated with FEV_1_, but only 47 proteins significantly associated with FEV_1_ after adjusting for multiple-testing (FDR p-value < 0.2). In the MESA-Lung cohort there were 330 SOMAmers representing 315 proteins that were nominally significant, with 8 and 7 respectively that remained FDR significant. Notably, leptin was FDR significant (adjusted p-value < 0.2) in all three cohorts and higher values of leptin were associated with lower decline in FEV_1_ over the 5 year follow up (**Figure 2B**). Other protective proteins included growth hormone receptor (GHR) and fatty acid binding protein 3 (FABP3). GHR was FDR significant in the two COPD-enriched cohorts. Insulin growth factor binding protein-2 (IGFBP-2) is an example of an opposite relationship in that higher levels of the IGFBP-2 were associated with faster FEV_1_ decline (risk proteins).

**Figure 2:**
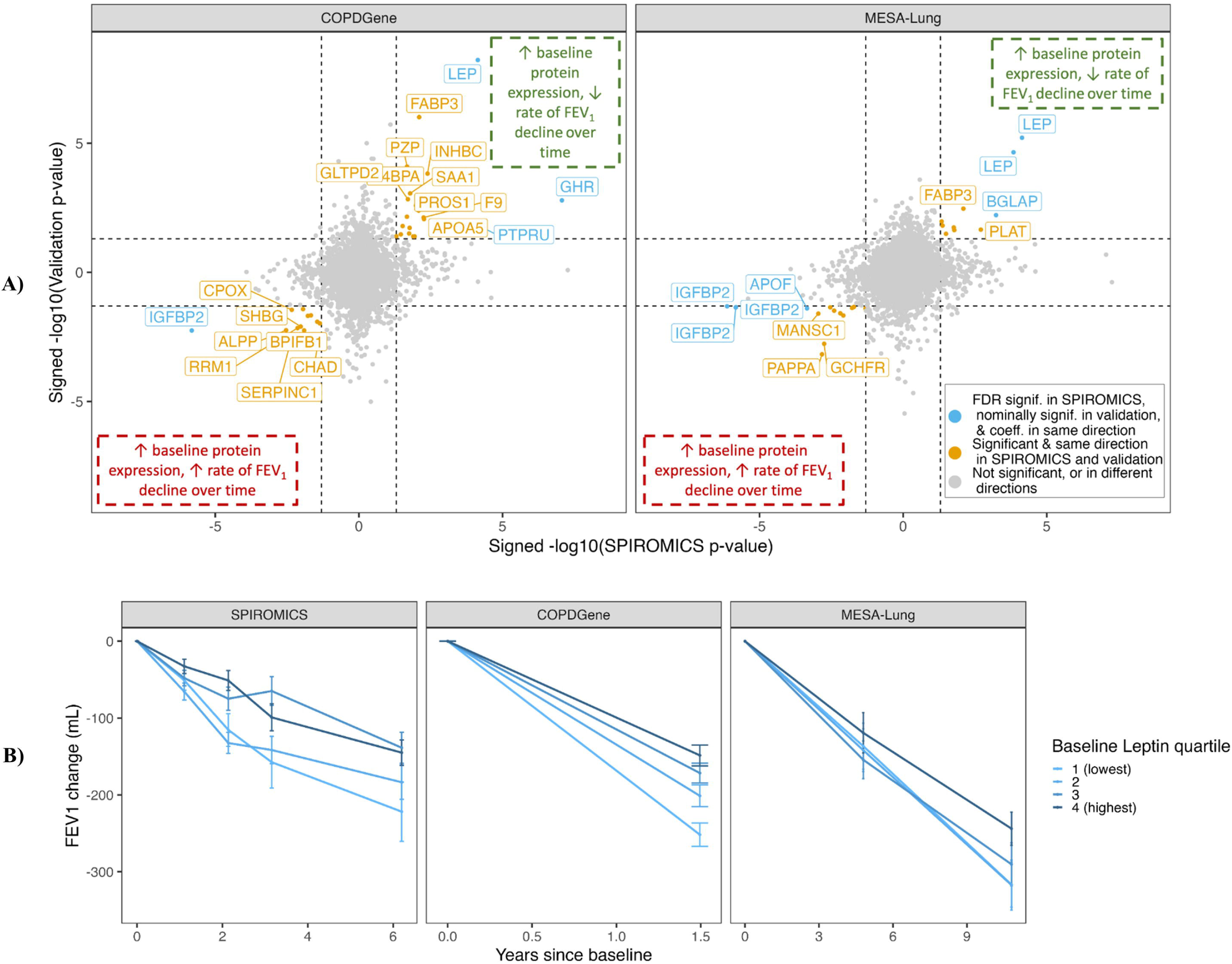
Baseline proteins associated with FEV_1_ change over time in SPIROMICS, COPDGene, and MESA-Lung. (A) The x-axes of each panel display the signed -log_10_(p-value) (sign of beta estimate * -log_10_(p-value)) from our discovery cohort, SPIROMICS. The y-axes show the signed -log_10_(p-value) from our validation cohorts, COPDGene and MESA-Lung. Proteins highlighted in orange overlapped between our discovery cohort and one of our validation cohorts (nominal p-value < 0.05 and beta estimate in same direction). Proteins highlighted in blue were FDR significant (adjusted p-value < 0.2) in discovery and nominally significant (p-value < 0.05) in validation, with beta estimate in the same direction. Points are labeled with Entrez Gene symbol, see Supp. Table 1 for corresponding protein target name and additional information. (B) Subjects in each cohort were stratified into 4 quartiles based on their baseline leptin expression, with group 1 having the lowest baseline leptin and group 4 having the highest. The average change in FEV_1_ (post-bronchodilator for SPIROMICS and COPDGene, pre-bronchodilator for MESA-Lung) from baseline was calculated for each quartile group at each visit. Baseline leptin is predictive of rate of FEV_1_ change.

### Proteins which improve model fit for a multi-variate COPD progression model

Next, we examined which of the seven proteins with nominal significance in all three cohorts could improve FEV_1_ progression model fit compared to using only clinical and demographic characteristics (clinical model). Most proteins individually improved AIC, BIC, and marginal R^2^ in both SPIROMICS and COPDGene (**<u>Table E2</u>**). Using backward selection, we found a combination of 3 proteins (placental alkaline phosphatase (ALPP), IGFBP-2, and leptin) best improved the clinical model. ALPP, a well-known biomarker of smoking, improved modeling of FEV_1_ progression even with adjustment for current smoking, suggesting that treating smoking as a binary variable may incompletely capture the effect of continued smoking on progression of COPD. In the SPIROMICS cohort we found only a weak relationship between ALPP and number of cigarettes smoked per day or urinary cotinine level in smokers, thus it’s not clear that there is significantly better modeling with smoking history. Alternatively, ALPP is also an oncoprotein and can bind viruses such as Zika. Thus, there may be functions of ALPP not related to smoking.

### A model estimating annualized decline in FEV_1_

To assess the clinical value of the progression model created using SPIROMICS data, we used this model to calculate predicted change in COPDGene subjects and compared this to observed annualized change in FEV_1_ (**Figure 3**). This independent clinical/protein progression model validation suggests that our model performs well in identifying subjects at high and low risk of progression. For instance, the first decile in SPIROMICS and COPDGene had an observed median FEV_1_ mL/year change of -127.226 and -107.132 respectively, while the tenth decile had 42.213 and 37.705 **(<u>Table E2</u>).**

**Figure 3:**
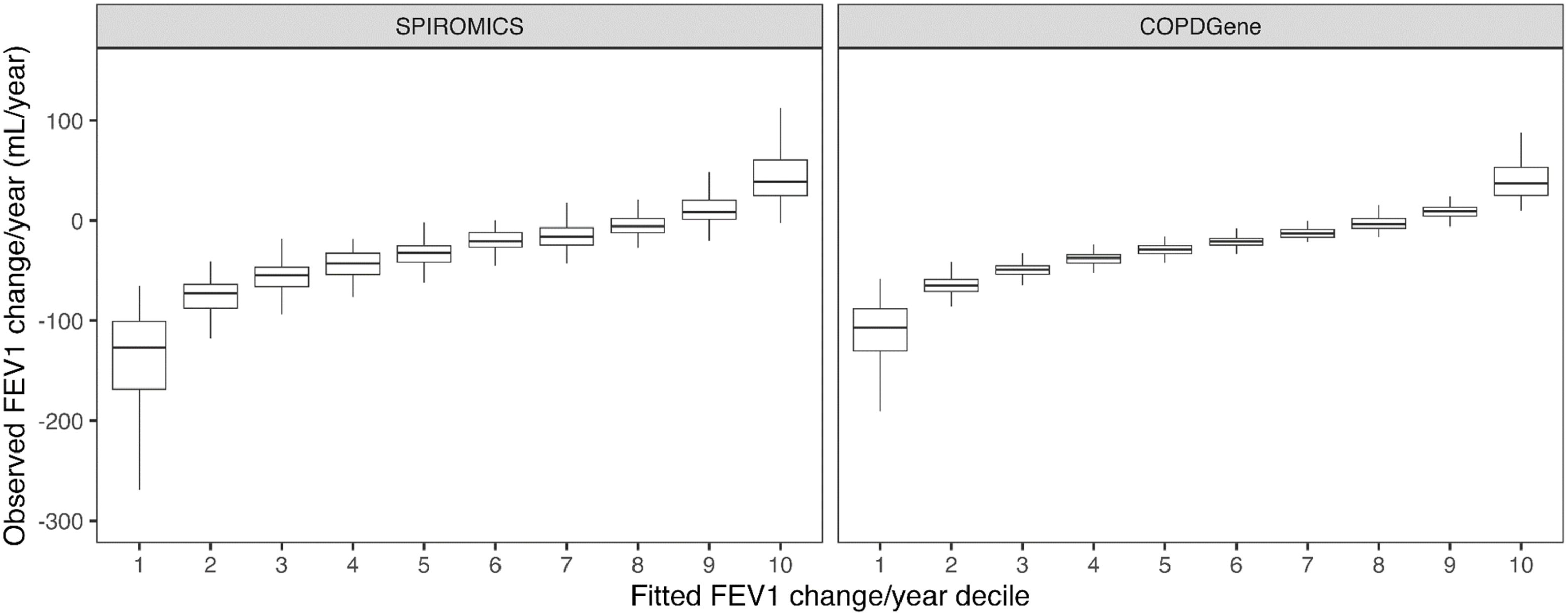
Predicted FEV_1_ change per year by composite prognostic model versus observed FEV_1_ change per year for COPD enriched cohorts (SPIROMICS and COPDGene). The plot is limited to the last follow-up visit at least 3 years after baseline for each subject.

### Dynamic Protein Changes Associated with Changes in FEV_1_

Many SPIROMICS and COPDGene subjects had multiple visits with proteomic assessments **(<u>Table E6</u>)**. Thus, we could evaluate whether there are protein biomarkers whose changes were associated with FEV_1_ (dynamic or disease activity biomarkers). In SPIROMICS, longitudinal changes in FEV_1_ decline were associated with changes in 686 proteins (717 SOMAmers) at p-value < 0.05 and 155 proteins (163 SOMAmers) at FDR p-value < 0.2. 69 proteins (71 SOMAmers) were nominally significant in both cohorts, but none were FDR significant in both cohorts. Twelve of these 69 proteins were FDR significant in SPIROMICS and nominally significant in COPDGene **(Figure 4, Table E1).** Some of the stronger associations were for Phosphoglycerate mutase 1 & 2 (PGAM1 & PGAM 2), which play key roles in glycolysis, and Glutamyl aminopeptidase (ENPEP), which cleaves N-terminal aspartate or glutamate, thereby changing protein functions for diverse functions such as capillary angiogenesis.

**Figure 4:**
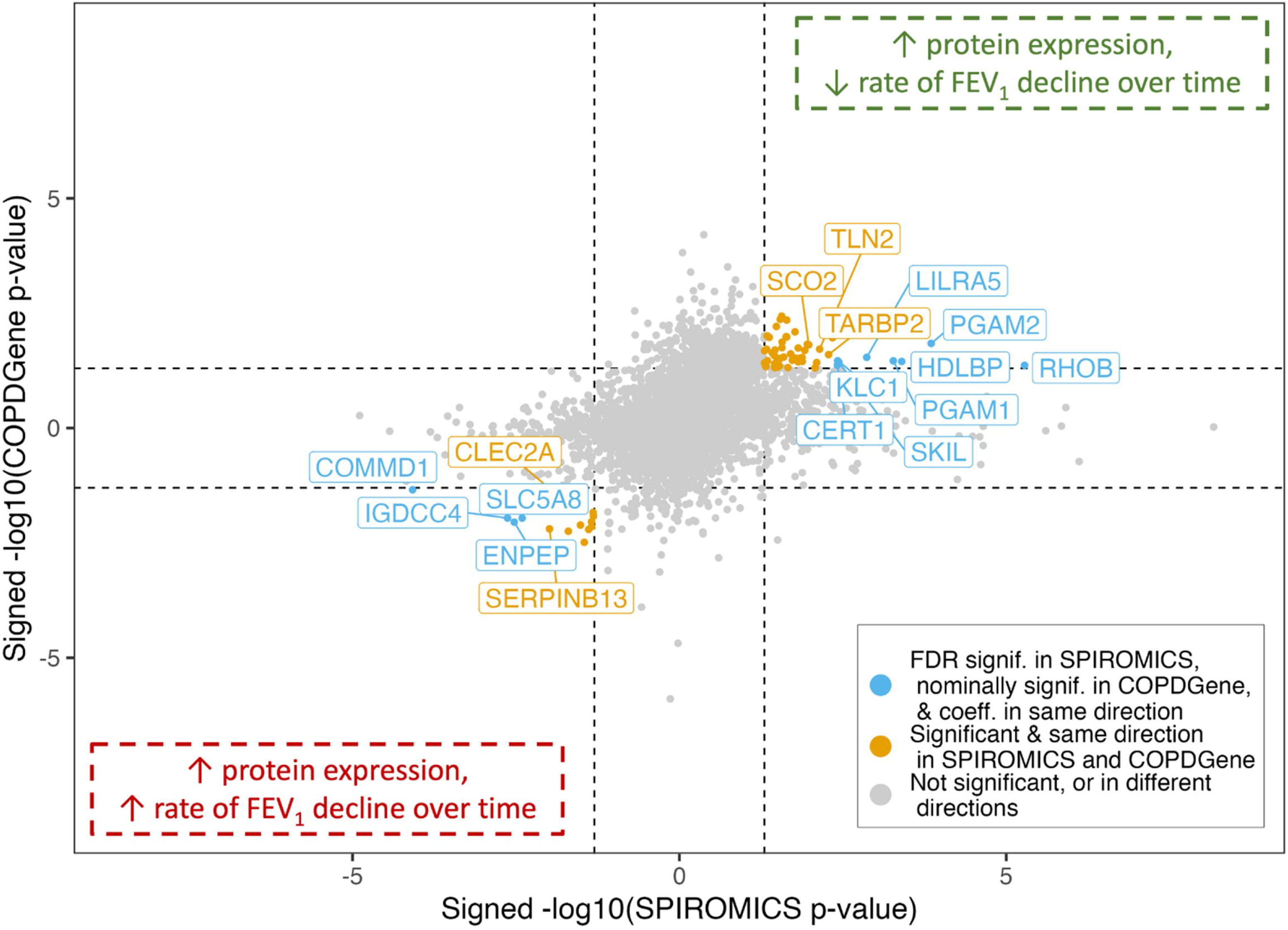
Proteins dynamically associated with FEV_1_ over time in SPIROMICS and COPDGene. The axes display the signed -log_10_(p-value) (sign of beta estimate * - log_10_(p-value)) from our discovery cohort, SPIROMICS, and our validation cohort, COPDGene. 72 proteins were identified that overlapped between two cohorts changing in the same direction (nominal p-value < 0.05) highlighted in orange. 12 of those proteins highlighted in blue were FDR significant (adjusted p-value < 0.2) in SPIROMICS. Points are labeled with Entrez Gene symbol, see Supp. Table 4 for corresponding protein target name and additional information.

### GSEA identifies similarities between SPIROMICS & COPDGene in protein expression associated with FEV_1_ decline

To determine which pathways associated with COPD progression were in common between COPDGene and SPIROMICS, we used gene set enrichment analysis (GSEA) to find overlapping proteins and then used Metascape to identify pathways for both the prognostic and dynamical models (**Figure 5**). For the prognostic and dynamic models, there were 70 (adjusted p-value < 0.05) and 128 (adjusted p-value < 0.05) proteins that were significantly and concordantly enriched (**Figure 5**, **<u>Table E5</u>**). The most significant pathway in the prognostic model of COPD progression was the complement system and the most significant for the dynamic model was the PID AVB3 OPN pathway, which represents osteopontin mediated events (**Figure E2**). Although the peptide hormone metabolism pathway was common between the prognostic and dynamic models, the majority of functional categories were distinct, suggesting that the protein pathways that represent prognostic value for predicting future changes in FEV_1_ are different from the pathways that change with disease activity (**Figure E2**).

**Figure 5:**
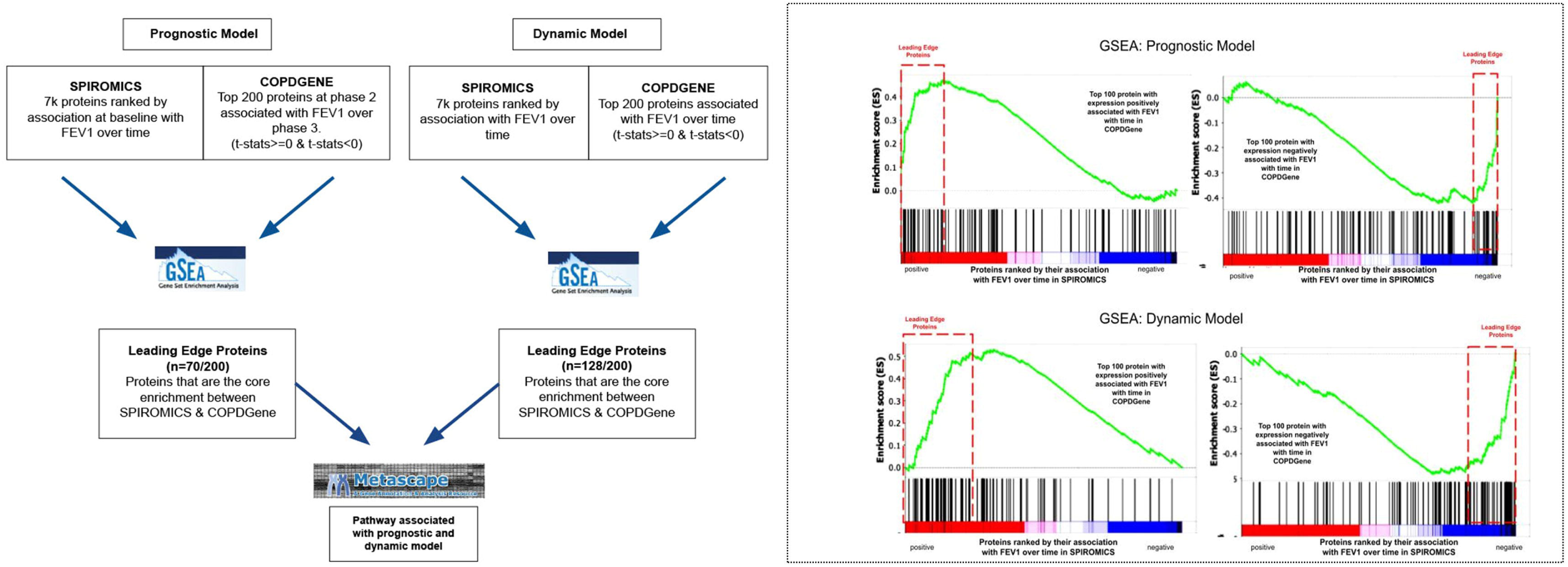
Enrichment between COPD datasets for prognostic and dynamic biomarkers in response to change in FEV_1_ over time. (A) Figure shows the strategy flow diagram that was used to identify proteins that are most enriched between the SPIROMICS and COPDGene data sets for the Prognostic and Dynamic model separately. The leading-edge proteins were then passed to functional annotation tool, Metascape, to identify the biological similarities between the two models. B) The relationship between the effect of protein expression on FEV_1_ overtime in the COPD

## DISCUSSION

This is one of the largest studies leveraging high-throughput proteomic profiling to identify prognostic and dynamic biomarkers associated with longitudinal changes in FEV_1_ in three large, well-phenotyped cohorts (COPDGene, SPIROMICS and MESA-Lung). Our study validated leptin as a prognostic biomarker with higher baseline leptin levels associated with slower FEV_1_ decline across all cohorts. Higher baseline growth hormone receptor (GHR) was also associated with slower rates of decline, but only in the COPD specific cohorts. Additionally, there were dozens of novel prognostic biomarkers associated with change in FEV_1_ at nominal significance across both COPD cohorts. Many of these individual biomarkers only partially explained spirometric progression and a combination of 3 proteins performed best in predicting FEV_1_ decline. This finding aligns with the concept that COPD progression results from multiple disparate pathologic processes, and further supports the notion that multiple biomarkers are needed to predict disease progression (9).

We are among the first to report the association of higher leptin concentrations with a slower rate of change in FEV_1_ in a model adjusting for BMI and FVC (11). Previous studies found leptin inversely correlated with baseline FEV_1_. However, these studies were limited to population-based cohorts or not powered to show an association between leptin and longitudinal change in FEV_1_ (12–14). Other small studies reported inconsistent associations between leptin and lung function (15–17). One hypothesis in COPD is that leptin, although a known proinflammatory cytokine, contributes to antimicrobial defenses that are crucially involved in protection against early small airways disease (18). Additionally, leptin is essential to leukocyte function and genetic deficiency of leptin impairs host defenses against respiratory tract infections in humans and mice (19–21). Leptin is highly associated with obesity (high BMI), which is in turn associated with diseases such as diabetes and cardiovascular disease. Conversely, low BMI in the COPD population is associated with a 3-fold increase in mortality (22). This suggests that high leptin levels could be protective for lung function decline, but excess leptin could be deleterious promoting inflammation (obesity paradox) (23).

GHR is another energy/growth protein which exhibited strong associations with COPD progression in COPD enriched cohorts (COPDGene and SPIROMICS), but not in the population cohort (MESA-Lung). GHR, like leptin receptor, is a type 1 cytokine receptor. Intriguingly, leptin signaling may directly influence growth hormone production through the hypothalamus pituitary access, such that low leptin levels decrease growth hormone secretion leading to down regulation of growth hormone receptor via growth hormone stimulated processes (24). We also identified insulin-like growth factor binding protein-2 (IGFBP-2) as a prognostic biomarker that was FDR significant in SPIROMICS and nominally significant in COPDGene and MESA-Lung. IGFBP-2 regulates the bioavailability of insulin like growth factor (25). Further research is needed to understand how higher leptin levels are protective while higher levels of IGFBP-2 are associated with faster progression.

We found that a composite model (including baseline clinical, demographic, and multiple proteins) predicting FEV_1_ over time performed better than models using only single or no proteins. These observations are supported by Zemans et al. who concluded that models containing multiple proteins improve predictive value of COPD outcomes compared to clinical variables or individual proteins (9). However, this study used AIC to assess model improvement and not more strict FDR criteria for significance.

A unique feature of this study is the use of two different approaches to identify proteins predictive of FEV_1_ decline and dynamically modulated with FEV_1_ decline in multiple large COPD cohorts. Our dynamic model identified 12 proteins that significantly change with longitudinal FEV_1_ change in SPIROMICS with nominal significance in COPDGene. All of these proteins have novel associations with COPD, except glutamyl aminopeptidase (ENPEP) and high-density lipoprotein binding protein (HDLBP) (26–28). We identified several pathways that were significantly prognostic or dynamic, suggesting they may be relevant in both early disease and continue to play a role in disease progression. The osteopontin-mediated pathway was the most significant dynamically changing pathway for both SPIROMICS and COPDGene cohorts. Osteopontin, a crucial lung matrix glycoprotein and cytokine, plays a pivotal role in immune cell recruitment, tissue repair, angiogenesis, and remodeling (29). Additionally, the complement pathway ranked high in enrichment among leading-edge proteins in the prognostic model. Several complement factors have been implicated in COPD’s development, progression, and exacerbation, contributing to the inflammatory response within the lungs (30). Furthermore, some complement factors have been observed to be dysregulated in COPD (40). Modulating complement pathways is currently being explored as a therapeutic approach for various diseases, including conditions such as Paroxysmal Nocturnal Hemoglobinuria (PNH) and kidney disease. Overall, this promising avenue of research holds the potential for further exploration in the context of COPD.

Strengths of our study include intra-individual repeated measures design, inclusion of Black Americans, and large sample size from three different observational cohorts, which provides more statistical power compared to cross-sectional and independently analyzed observational studies. Nevertheless, our study findings may have been limited by clinical and disease heterogeneity across cohorts. Another potential limitation included the differences in SomaScan platform (5k vs 7k profiling platforms), but we used GSEA for comparison across datasets to address this issue. Depending on the context of use, future work to advance biomarker candidates into clinical practice or clinical trials, may require confirmatory standard clinical immunoassays. Future analysis could focus on endotyping of COPD patients using proteomic profiling from these cohorts to reduce the disease heterogeneity and help identify biomarkers to specific COPD subpopulations.

In summary, utilizing three large observational cohorts, we have identified protein biomarkers such as leptin that predict and dynamically change with FEV_1_ over time and that some of the COPD-associated immune response in the serum may be mediated by leptin. The known biological links between leptin and other leading proteins identified in this analysis (GHR and IGFBP-2) suggest that leptin may play a central mechanistic role in modulating spirometric decline in individuals with COPD.

## Data Availability

All data produced in the present study are available upon reasonable request to the authors

## SPIROMICS

The authors thank the SPIROMICS participants and participating physicians, investigators, study coordinators, and staff for making this research possible. More information about the study and how to access SPIROMICS data is available at www.spiromics.org. The authors would like to acknowledge the University of North Carolina at Chapel Hill BioSpecimen Processing Facility (http://bsp.web.unc.edu/) and Alexis Lab (https://www.med.unc.edu/cemalb/facultyresearch/alexislab/) for sample processing, storage, and sample disbursements. We would like to acknowledge the following current and former investigators of the SPIROMICS sites and reading centers: Neil E Alexis, MD; Wayne H Anderson, PhD; Mehrdad Arjomandi, MD; Igor Barjaktarevic, MD, PhD; R Graham Barr, MD, DrPH; Patricia Basta, PhD; Lori A Bateman, MS; Christina Bellinger, MD; Surya P Bhatt, MD; Eugene R Bleecker, MD; Richard C Boucher, MD; Russell P Bowler, MD, PhD; Russell G Buhr, MD, PhD; Stephanie A Christenson, MD; Alejandro P Comellas, MD; Christopher B Cooper, MD, PhD; David J Couper, PhD; Gerard J Criner, MD; Ronald G Crystal, MD; Jeffrey L Curtis, MD; Claire M Doerschuk, MD; Mark T Dransfield, MD; M Bradley Drummond, MD; Christine M Freeman, PhD; Craig Galban, PhD; Katherine Gershner, DO; MeiLan K Han, MD, MS; Nadia N Hansel, MD, MPH; Annette T Hastie, PhD; Eric A Hoffman, PhD; Yvonne J Huang, MD; Robert J Kaner, MD; Richard E Kanner, MD; Mehmet Kesimer, PhD; Eric C Kleerup, MD; Jerry A Krishnan, MD, PhD; Wassim W Labaki, MD; Lisa M LaVange, PhD; Stephen C Lazarus, MD; Fernando J Martinez, MD, MS; Merry-Lynn McDonald, PhD; Deborah A Meyers, PhD; Wendy C Moore, MD; John D Newell Jr, MD; Elizabeth C Oelsner, MD, MPH; Jill Ohar, MD; Wanda K O’Neal, PhD; Victor E Ortega, MD, PhD; Robert Paine, III, MD; Laura Paulin, MD, MHS; Stephen P Peters, MD, PhD; Cheryl Pirozzi, MD; Nirupama Putcha, MD, MHS; Sanjeev Raman, MBBS, MD; Stephen I Rennard, MD; Donald P Tashkin, MD; J Michael Wells, MD; Robert A Wise, MD; and Prescott G Woodruff, MD, MPH. The project officers from the Lung Division of the National Heart, Lung, and Blood Institute were Lisa Postow, PhD, and Lisa Viviano, BSN.

## COPDGene

COPDGene® Investigators – Core Units: Administrative Center: James D. Crapo, MD (PI); Edwin K. Silverman, MD, PhD (PI); Barry J. Make, MD; Elizabeth A. Regan, MD, PhD; Genetic Analysis Center: Terri Beaty, PhD; Ferdouse Begum, PhD; Peter J. Castaldi, MD, MSc; Michael Cho, MD; Dawn L. DeMeo, MD, MPH; Adel R. Boueiz, MD; Marilyn G. Foreman, MD, MS; Eitan Halper-Stromberg; Lystra P. Hayden, MD, MMSc; Craig P. Hersh, MD, MPH; Jacqueline Hetmanski, MS, MPH; Brian D. Hobbs, MD; John E. Hokanson, MPH, PhD; Nan Laird, PhD; Christoph Lange, PhD; Sharon M. Lutz, PhD; Merry-Lynn McDonald, PhD; Margaret M. Parker, PhD; Dmitry Prokopenko, Ph.D; Dandi Qiao, PhD; Elizabeth A. Regan, MD, PhD; Phuwanat Sakornsakolpat, MD; Edwin K. Silverman, MD, PhD; Emily S. Wan, MD; Sungho Won, PhD; Imaging Center: Juan Pablo Centeno; Jean-Paul Charbonnier, PhD; Harvey O. Coxson, PhD; Craig J. Galban, PhD; MeiLan K. Han, MD, MS; Eric A. Hoffman, Stephen Humphries, PhD; Francine L. Jacobson, MD, MPH; Philip F. Judy, PhD; Ella A. Kazerooni, MD; Alex Kluiber; David A. Lynch, MB; Pietro Nardelli, PhD; John D. Newell, Jr., MD; Aleena Notary; Andrea Oh, MD; Elizabeth A. Regan, MD, PhD; James C. Ross, PhD; Raul San Jose Estepar, PhD; Joyce Schroeder, MD; Jered Sieren; Berend C. Stoel, PhD; Juerg Tschirren, PhD; Edwin Van Beek, MD, PhD; Bram van Ginneken, PhD; Eva van Rikxoort, PhD; Gonzalo Vegas Sanchez-Ferrero, PhD; Lucas Veitel; George R. Washko, MD; Carla G. Wilson, MS; PFT QA Center, Salt Lake City, UT: Robert Jensen, PhD; Data Coordinating Center and Biostatistics, National Jewish Health, Denver, CO: Douglas Everett, PhD; Jim Crooks, PhD; Katherine A. Pratte, PhD; Matt Strand, PhD; Carla G. Wilson, MS; Epidemiology Core, University of Colorado Anschutz Medical Campus, Aurora, CO: John E. Hokanson, MPH, PhD; Gregory Kinney, MPH, PhD; Sharon M. Lutz, PhD; Kendra A. Young, PhD; Mortality Adjudication Core: Surya P. Bhatt, MD; Jessica Bon, MD; Alejandro A. Diaz, MD, MPH; MeiLan K. Han, MD, MS; Barry Make, MD; Susan Murray, ScD; Elizabeth Regan, MD; Xavier Soler, MD; Carla G. Wilson, MS; Biomarker Core: Russell P. Bowler, MD, PhD; Katerina Kechris, PhD; Farnoush Banaei-Kashani, Ph.D

## MESA

The authors thank the other MESA-Lung investigators, the staff, and the participants of the MESA study for their valuable contributions. A full list of participating MESA investigators and institutions can be found at http://www.mesa-nhlbi.org [mesa-nhlbi.org]. This paper has been reviewed and approved by the MESA Publications and Presentations Committee.

## Author Contributions

Shared-first authorship is indicated by an asterisk (*) amongst the first four authors.

## AT A GLANCE COMMENTARY

### Scientific Knowledge on the Subject

Cross sectional studies have identified COPD protein biomarkers and biologic pathways that are associated with spirometric measures of airflow obstruction and reduced forced expiratory volume at one second (FEV_1_); a smaller number of studies have found protein biomarkers that are associated with accelerated decline in FEV_1_. However, large scale COPD prognostic proteomic studies with replication across diverse cohorts and studies relating progression of airflow obstruction with dynamic changes in protein biomarkers are lacking.

### What This Study Adds to the Field

In three large longitudinal studies that include non-Hispanic White and Black Americans, we identified multiple replicated protein biomarkers associated with COPD progression measured by spirometry. The strongest association with COPD progression was with leptin, for which higher plasma levels were associated with less COPD progression.

## Figure legends

**Supplemental Figure El:**
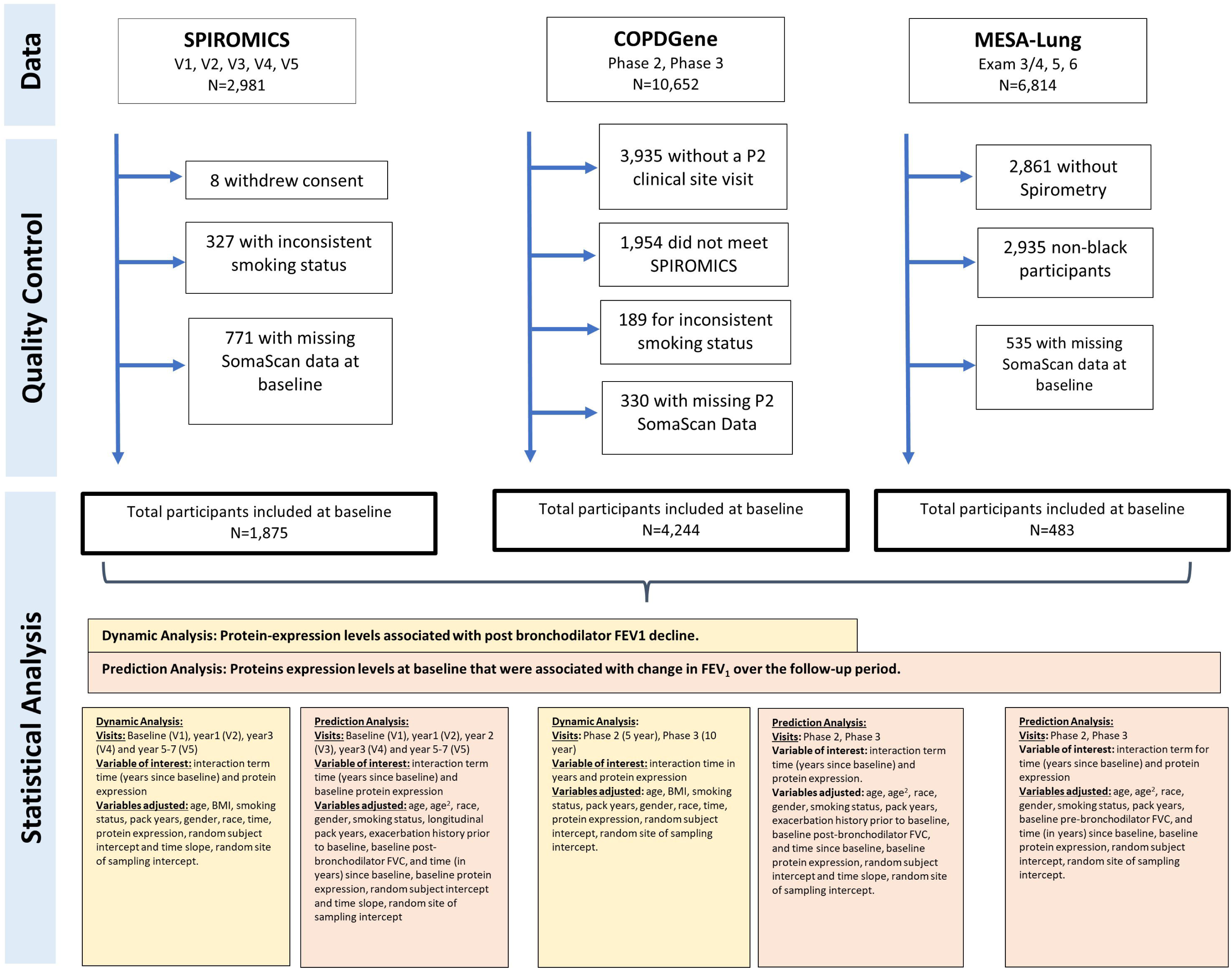
Study design flow chart.

**Supplemental Figure E2:**
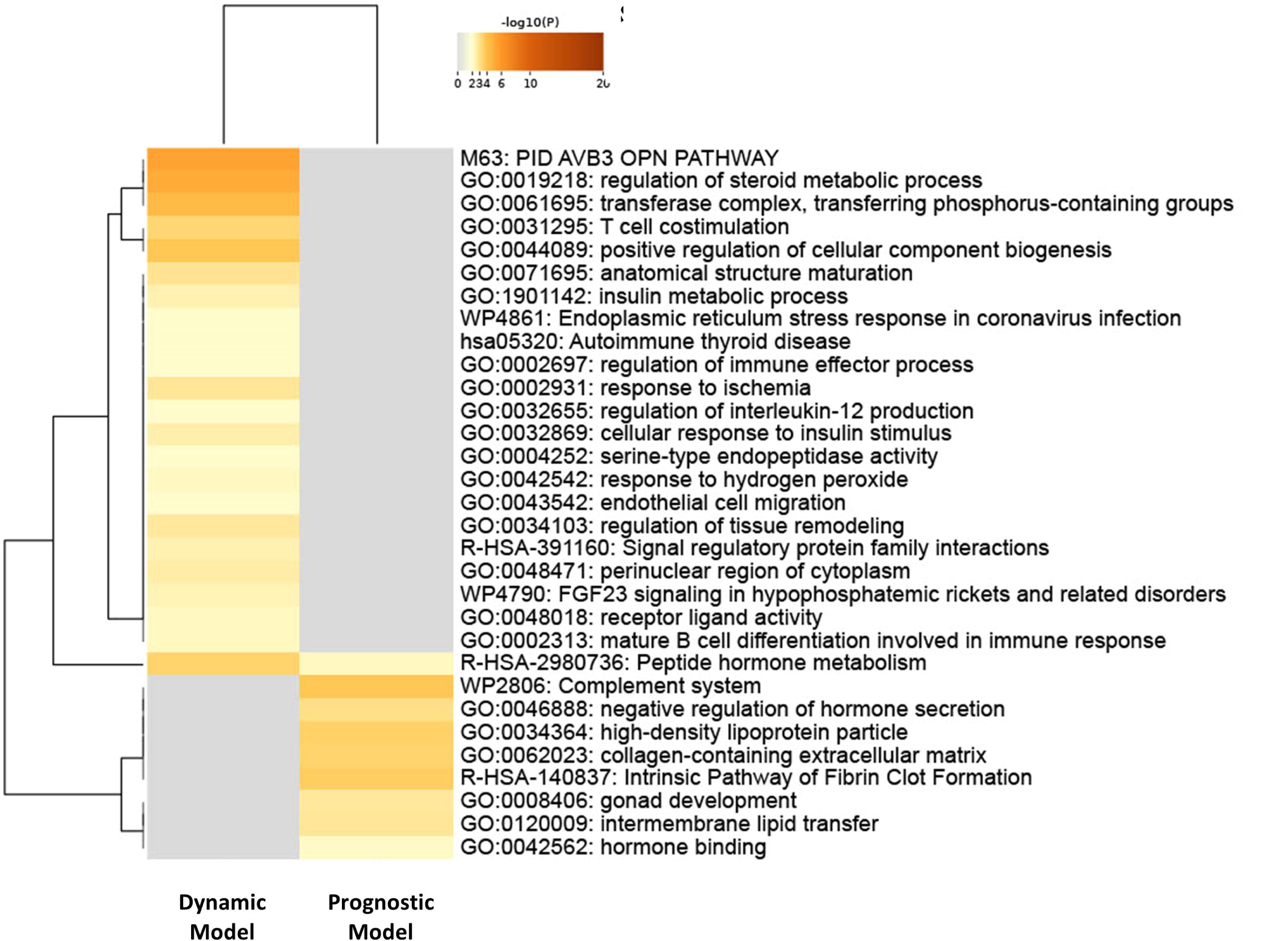
Functional Enrichment: of markers of FEV_1_ over time.

**Supplemental Figure E3:**
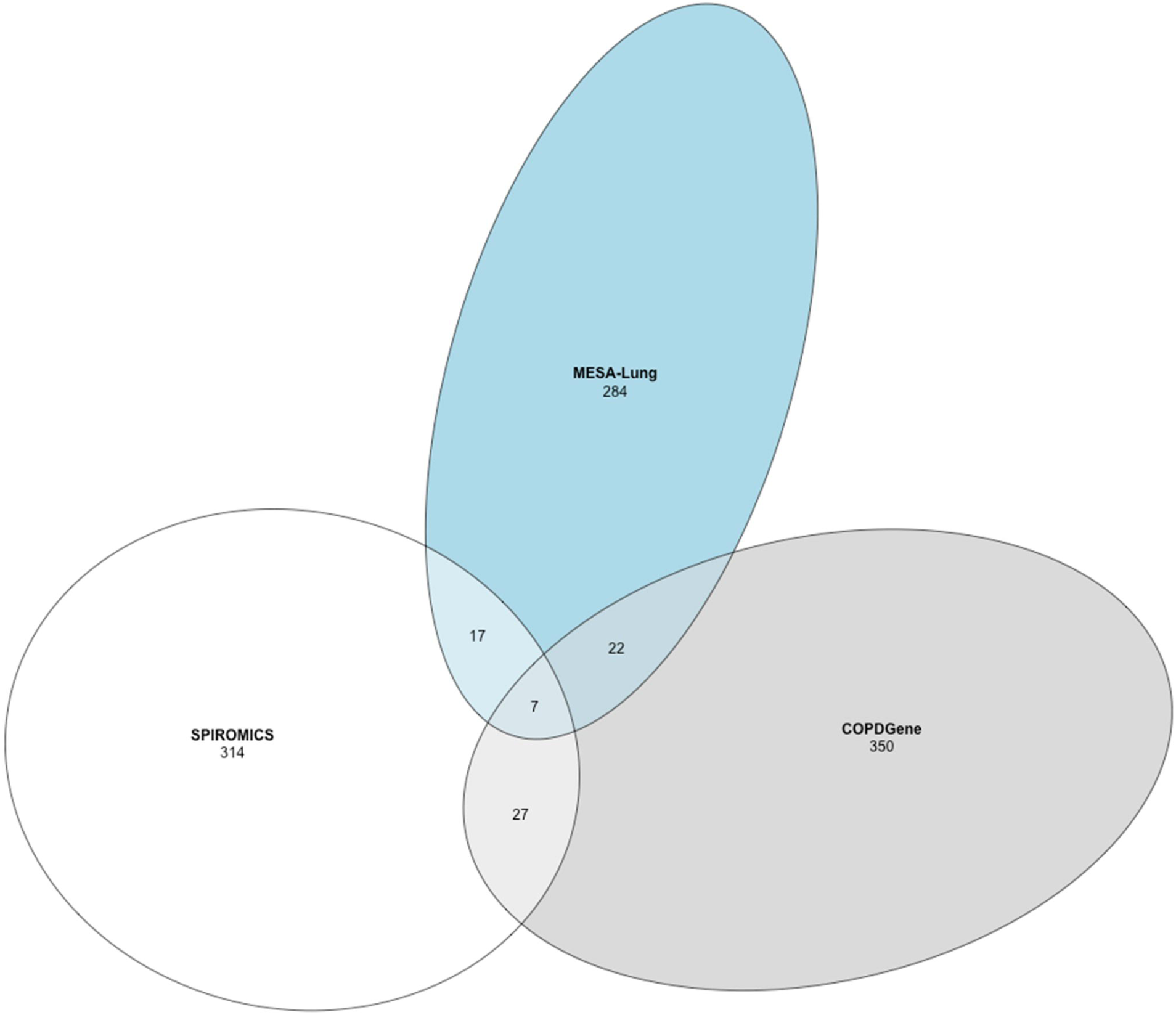
Proteins overlapping in each cohort by nominal significance and with coefficient estimates in the same direction from the prognostic model.

