## supplemental text for "Dynamic and prognostic proteomic associations with FEV_1_ decline in chronic obstructive pulmonary disease"

Cohort Descriptions

SPIROMICS (ClinialTrials.gov Identifier: NCT01969344), used 12 clinical centers to enroll individuals 40-80 years old, all races, with and without COPD between 2010-2015. The study recruited n=200 never smokers (<1 pack year history) and n=2,788 current or former smokers with ≥ 20-pack year history. After initial visits, participants were asked to participate in annual follow-up visits for the first three years and a 5^th^ visit occurring approximately 5-7 years later. Participants underwent pre- and post-bronchodilator spirometry at all visits and blood draws at baseline, year 1, year 3 and a final visit that occurred approximately 5-7 years later. Additional details can be found in (1).

COPDGene (ClinialTrials.gov Identifier: NCT00608764), used 21 centers across the United States to enroll individuals between ages 45-80 years between 2008 to 2011. The study recruited non-Hispanic Whites and Blacks in a roughly 2:1 ratio enrolling n=454 never-smoking (<100 cigarettes in lifetime) and n=10,198 current and former smokers (≥10 pack year history) with and without COPD. Participants were invited for a 5-year and 10 year follow up. Participants had pre- and post-bronchodilator spirometry and blood draws at all visits. Additional details can be found in (2).

MESA (ClinialTrials.gov Identifier: NCT00005487), is a multicenter prospective cohort study designed to investigate the prevalence, correlates, and progression of subclinical cardiovascular disease in individuals without clinical cardiovascular disease (3, 4). MESA recruited 6,814 men and women ages 45-84 years old in 2000-2002 from six U.S. communities. MESA participants are White, Black, Hispanic, or Asian (mostly of Chinese origin). The MESA-Lung Study, an ancillary study that is the source of this analysis, enrolled n=3,965 MESA participants (of 4,484 selected) who were sampled randomly among those who consented and attended an examination in 2004-2006 (99%, 89%, and 91% of the MESA cohort, respectively). Proteomic data in MESA-Lung was available only for Black participants, of which 483 participants had both spirometry measurements and proteomic data. All measures were obtained at the MESA baseline examination, except for spirometry, which was performed during the MESA-Lung recruitment period.

Biomarker Panel

We assayed participant plasma using SomaScan (Somalogic, Inc., Boulder), which employs a pool of specific aptamers (SOMAmers) that bind to epitopes in the sample matrix, to provide relative abundances from a sample volume ≤60 µ (5). This study used both SomaScan v4.1 (SPIROMICS, MESA) and v4.0 (COPDGene). SomaScan v4.1 makes 7,288 SOMAmer measurements (“7k”) mapping to 6,401 Human Uniprot IDs. SomaScan v4.0 used 4,979 SOMAmers (“5k”) mapping 4,776 unique human proteins. SomaScan measurements are made on a custom Agilent oligo microarray that yields a Relative Fluorescence Unit (RFU) for each SOMAmer. Data received from Somalogic is subject to technical quality assessment by applying a standardized analysis anchored on library(arrayQualityMetrics). The SomaScan assay includes a 96-well plate format. Each plate for their core matrices contains replicate controls of a) buffer only, b) stock calibrator matching submitted sample matrix, and c) QC samples of same matrix for evaluation of assay run conditions and normalization parameters. Both 96-well plate IDs and Agilent Scanner IDs are tracked and evaluated in the process. Individual submitted samples are evaluated by three outlier detection methods applied in arrayQualityMetrics, and samples with extreme D-statistic and corresponding MA plot are flagged. Along with SomaLogic’s outlier flagging of normalization deviants, these flags inform subsequent analytics once submitted samples are re-identified. Results were flagged and removed if they did not pass quality assurance (6, 7).

SomaLogic standardized the SomaScan® data per their protocol. Their normalization, calibration, and scaling consisted of: “within plate hybridization to control for variability across array signals, median signal normalization to control for technical variability of replicates within a run, plate scaling and calibration of SOMAmers to control for inter-assay variation between analytes and batch differences between plates. Median normalization to a reference using adaptive normalization by maximum likelihood is applied within dilution group to quality control replicates and individual samples to remove edge effects and technical variance (5). As described by SomaLogic, the adaptive normalization by maximum likelihood uses an adaptive procedure which censors analytes during normalization that fall beyond the expectations for a measurement from the distribution of SomaLogic’s population reference. “The scale factor is calculated by maximizing the probability that a sample’s RFU measurement came from the reference distribution. This is accomplished by using the information of the reference population variance for each analyte in addition to the reference distribution median.” All values were log transformed prior to analysis (8).

### *Clinical Definitions and Study Population*

All cohorts collected demographic information, smoking history, medical history, and respiratory symptoms using validated questionnaires. COPD was defined by post-bronchodilator FEV_1_ to FVC ratio < 0.70; in MESA-Lung, only pre-bronchodilator spirometry measurements were available in Exam 3 and thus used for Exams 3 through 6 to define COPD. All three cohorts used self-defined race, chosen from a limited number of categories. We refer to the selected choices of “Black” and/or “African American” as “Black” and where participants selected “White” and/or the historically used category “Caucasian” we refer to their race/ancestry as “White.” Since over 50 percent of the enrolled participants in SPIROMICS and COPDGene were non-Hispanic white and COPD disproportionately affects minorities, we included only black MESA participants.

All participants who consented to genetic analysis and had available proteomics results and spirometry for at least 1 visit were included **(**[**Figure 1**](https://olucdenver-my.sharepoint.com/:w:/g/personal/lisa_ruvuna_cuanschutz_edu/EWo69ebiwFNIvgm_SHZ2wIQBQs8fjIz2E1VGjQezC1vAog?e=EKendl)**).** To better compare between COPDGene and SPIROMICS, COPDGene subjects were categorized into the 4 SPIROMICS enrollment strata and excluded if they did not meet strata criteria. We minimized variability caused by smoking in SPIROMICS and COPDGene by analyzing subjects who maintained consistent smoking status throughout the study. Since MESA-Lung is a population cohort and had a smaller sample size, we did not limit to those who maintained consistent smoking status and we did not apply the SPIROMICS enrollment criteria.

After quality checks and data curation, the final datasets consisted of N=1,875 subjects for SPIROMICS, N=4,244 for COPDGene and N=483 for MESA **(**[**Figure 1**](https://olucdenver-my.sharepoint.com/:w:/g/personal/lisa_ruvuna_cuanschutz_edu/EWo69ebiwFNIvgm_SHZ2wIQBQs8fjIz2E1VGjQezC1vAog?e=EKendl)**).**

Statistical Analysis

### *Prognostic Model*

All analyses were performed in R. We used a linear mixed effects model to identify proteins at baseline associated with FEV_1_ over the follow-up period. We performed sensitivity analysis testing several clinical covariates and included those which improved the prediction accuracy of the model based on Akaike information criterion (AIC). These clinical covariates became our fixed effects which included age, age^2^, race, sex, smoking status, time-varying pack years, exacerbation history prior to baseline, baseline post-bronchodilator FVC, and time (in years) since baseline (first visit with SomaScan measurement) for SPIROMICS and COPDGene. Our main fixed effect of interest was the interaction of the protein with time. We included a random intercept and time slope at the subject level and a random intercept for each study center. The covariance matrix was unstructured. In MESA, the random slope was removed due to model convergence issues; the model is otherwise the same. We will refer to this model as a prognostic model as it identifies prognostic biomarkers.

We examined the protein*years interaction term p-values, as well as adjusted the p-values using the Benjamini-Hochberg method. A protein was considered nominally significant if the unadjusted p-value < 0.05, and FDR significant if the adjusted p-value was < 0.2. Using SPIROMICS as our discovery cohort, and COPDGene and MESA as our validation cohorts, we looked at the nominally significant proteins and the FDR significant proteins overlapping among the three cohorts. Additionally, we looked at proteins that were FDR significant in SPIROMICS (discovery cohort) and either FDR or nominally significant in one additional cohort (COPDGene or MESA). We also ensured the protein*years coefficient estimates were in the same direction across cohorts.

Assessing Performance of a Multi-Protein Prognostic Model

To determine if proteins of interest improved prediction of FEV1 using demographic and clinical characteristics in SPIROMICS, we used our prognostic model without the marker*years and marker coefficients as our base clinical model. We examined the Akaike information criteria (AIC), Bayesian information criteria (BIC), and marginal R2 when each protein was added to the base clinical model individually. We then evaluated the clinical model with all proteins of interest added, and used backward selection to arrive at a combination of proteins that best improves the clinical model. Specifically, we removed the least significant marker*years terms iteratively until all remaining marker*years terms had a p-value < 0.05. We then repeated this process for the marker main effect terms that did not have interaction terms in the model. To prevent collinearity, we first calculated the Pearson correlation between each pair of proteins and only used the most significant protein in backward selection if a pair had a correlation greater than 0.8. The backward selection process was done in our discovery cohort, SPIROMICS, and then the final selected model was tested in our validation cohort, COPDGene.

### *Dynamic Model*

To understand the association of protein expression kinetics with changes in lung function, we modeled post-bronchodilator FEV_1_ verse the interaction of longitudinal protein expression and time (in years) using a linear mixed effect model, independently in SPIROMICS and COPDGene. Covariates were selected based on their biological relevance while considering multi-collinearity between variables. We controlled for age, body mass index (BMI), smoking status, smoking pack-years, sex, and race. The same random effects were used as those in the prognostic model. The random slope was omitted in COPDGene as it was not estimable. We refer to this model as the dynamic model as it models dynamic protein changes associated with COPD progression. We examined the protein*years interaction term p-values and applied the same methodology as used in the prognostic model to identify nominal and FDR significant proteins within and overlapping between cohorts. This model was not assessed in MESA as only baseline SomaScan samples were available ([Figure 1](https://olucdenver-my.sharepoint.com/personal/lisa_ruvuna_cuanschutz_edu/Documents/manuscript%20progression%20COPD/Manuscript%20figures.docx?web=1)).

### *Gene Set Enrichment Analysis to identify proteins in common between COPD-enriched Cohorts*

We used Gene Set Enrichment Analysis (GSEA) (v4.2.3) to identify relationships at protein levels associated with changes in FEV_1_ over time in SPIROMICS and COPDGene (9). GSEA was performed for the two modeling approaches described previously (prognostic and dynamic). In SPIROMICS, proteins (7k assay) were ranked by their t-statistics of response to association with change in FEV_1_ over time (rank list); in COPDGene (5k assay), the top 200 proteins associated with FEV_1_ over time were used as a gene set for the GSEA analysis. Significant enrichment was defined by an FDR corrected p-value < 0.05. Most strongly associated proteins with FEV_1_ over time between the two cohorts (leading edge proteins) were used for further downstream analysis.

*Identification of significant pathways within and between COPD-enriched cohorts using Metascape*

We utilized the Metascape online tool (10) to perform functional enrichment analysis of the leading-edge proteins using the 5k SomaScan platform proteins as the background reference protein database. Metascape allowed us to filter and identify statistically enriched terms based on accumulative hypergeometric p-values and enrichment factors. Metascape hierarchically clustered these enriched terms into a tree based on Kappa-similarities among their gene memberships. Default settings were specified that used the gene ontology (GO) Biological Processes library to identify functional pathways, and only include pathways with a minimum of 3 and maximum of 500 genes. A p-value cutoff of 0.05 was used to identify statistically significant pathways, and a minimum enrichment of 1.5 was required for a pathway to be included. Metascape generated a hierarchal clustered heatmap of -log_10_ p-values of pathways enriched that were either common or unique to each of the GSEA leading edge proteins.

8. Dubin RF, Deo R, Ren Y, Lee H, Shou H, Feldman H, *et al*. Analytical and Biological Variability of a Commercial Modified Aptamer Assay in Plasma Samples of Patients with Chronic Kidney Disease.
*J Appl Lab Med* 2023; 8: 491-503.

9. Subramanian A, Tamayo P, Mootha VK, Mukherjee S, Ebert BL, Gillette MA, *et al*. Gene set enrichment analysis: a knowledge-based approach for interpreting genome-wide expression profiles. *Proc Natl Acad Sci U S A* 2005; 102: 15545-15550.

10. Zhou Y, Zhou B, Pache L, Chang M, Khodabakhshi AH, Tanaseichuk O, *et al*. Metascape provides a biologist-oriented resource for the analysis of systems-level datasets. *Nat Commun* 2019; 10: 1523.
